# Immunogenicity and reactogenicity of booster vaccinations after Ad26.COV2.S priming

**DOI:** 10.1101/2021.10.18.21264979

**Authors:** R.S.G. Sablerolles, W.J.R. Rietdijk, A. Goorhuis, D.F. Postma, L.G. Visser, D. Geers, K.S. Schmitz, H.M. Garcia Garrido, M.P.G. Koopmans, V.A.S.H. Dalm, N.A. Kootstra, A.L.W. Huckriede, M. Lafeber, D. van Baarle, C.H. GeurtsvanKessel, R.D. de Vries, P.H.M. van der Kuy, on behalf of the SWITCH research group

## Abstract

**Background:** In face of the developing COVID-19 pandemic with a need for rapid and practical vaccination strategies, Ad26.COV2.S was approved as single shot immunization regimen. While effective against severe COVID-19, Ad26.COV2.S vaccination induces lower SARS-CoV-2-specific antibody levels compared to its mRNA-based counterparts. To support decision making on the need for booster vaccinations in Ad26.COV2.S-primed individuals, we assessed the immunogenicity and reactogenicity of homologous and heterologous booster vaccinations in Ad26.COV2.S-primed health care workers (HCWs).

**Methods:** The SWITCH trial is a single-(participant)-blinded, multi-center, randomized controlled trial among 434 HCWs who received a single Ad26.COV2.S vaccination. HCWs were randomized to no boost, Ad26.COV2.S boost, mRNA-1273 boost, or BNT162b2 boost. We assessed the level of SARS-CoV-2-specific binding antibodies, neutralizing antibodies against infectious virus, SARS-CoV-2-specific T-cell responses, and reactogenicity.

**Results:** Homologous and heterologous booster vaccinations resulted in an increase in SARS-CoV-2-specific binding antibodies, neutralizing antibodies and T-cell responses when compared to single Ad26.COV.2.S vaccination. In comparison with the homologous boost, the increase was significantly larger in heterologous regimens with the mRNA-based vaccines. mRNA-1273 boosting was most immunogenic, associated with higher reactogenicity. Only mild to moderate local and systemic reactions were observed on the first two days following booster.

**Conclusions:** Boosting of Ad26.COV2.S-primed HCWs was well-tolerated and immunogenic. Strongest responses were detected after boosting with mRNA-based vaccines. Based on our data, efficacy on infection and transmission of boosters is expected. In addition to efficacy, decision making on boost vaccinations should include timing, target population, level of SARS CoV-2 circulation, and the global inequity in vaccine access.

**Trial registration:** Funded by ZonMW (10430072110001); ClinicalTrials.gov number, NCT04927936.

## Introduction

Four COVID-19 vaccines are currently authorized for use in the European Union: two mRNA-based (BNT162b2 [‘Comirnaty’, Pfizer-BioNTech], mRNA-1273 [‘Spikevax’, Moderna]), and two adenovirus vector-based (ChAdOx1 nCoV-19 [‘Vaxzevria’, AstraZeneca], and Ad26.COV2.S [‘Janssen’]). Immunization with these vaccines prevents mild to severe COVID-19 with high efficacy ^1-4^. BNT162b2, mRNA-1273 and ChAdOx1 nCoV-19 are administered in homologous prime-boost regimens with different intervals, whereas Ad26.COV2.S is intended as a single-dose. The durability of protection and potential need for boosts is under continuous assessment.

The single-dose regimen and favorable storage conditions of Ad26.COV2.S provides major advantages in deployment ^3^. Additionally, both humoral and cellular immune responses are effectively induced, and persist up to 8 months after Ad26.COV2.S vaccination ^5,6^. However, in head-to-head comparisons, the mRNA-based vaccines induce higher levels of spike (S)-specific antibodies, and are superior to Ad26.COV2.S in vaccine efficacy (VE) against infection and mild disease ^7,8^. Notably, the difference in VE against hospitalization after Ad26.COV2.S or mRNA-based vaccination is smaller, probably in part attributed to immunological correlates like T-cell responses. Recent studies on homologous Ad26.COV2.S booster vaccination at 56 or 180 days after the primary vaccination showed an increase in binding antibody levels ^6^ but did not study effects on T-cell immunity, whereas this may play an important role in protecting against severe disease.

‘Mixing & matching’ different COVID-19 vaccines has the benefit of enhanced flexibility of vaccination campaigns ^9^, and the potential induction of broader immune responses ^10-14^. Heterologous vaccination regimens with ChAdOx1 nCoV-19 and BNT162b2 were previously demonstrated to be safe and to induce comparable or even superior immune responses than homologous boosting ^15-18^. Complete immunological and safety assessments on the effect of mRNA-boosters in Ad26.COV2.S-primed individuals have not yet been performed ^19^, but are highly relevant as millions of individuals have been immunized with this vaccine.

To support decision making regarding booster vaccinations in Ad26.COV2.S-primed individuals, we performed a head-to-head comparison in Ad26.COV2.S-primed HCWs of a homologous booster vaccination and heterologous boosting with mRNA-based vaccines BNT162b2 or mRNA-1273. A complete immunological assessment was performed, including the measurement of neutralizing antibodies and SARS-CoV-2-specific T-cell responses.

## Methods

### Ethical statement

The SWITCH trial is a single-(participant)-blinded, multi-center, randomized controlled trial among 434 healthy health care workers (HCWs) performed in four academic hospitals in the Netherlands (Amsterdam University Medical Center, Erasmus University Medical Center, Leiden University Medical Center, and University Medical Center Groningen) according to the previously described protocol ^9^. The trial adheres to the principles of the Declaration of Helsinki and was approved by the Medical Research Ethics Committee from Erasmus Medical Center (MEC 2021-0132) and the local review boards of the other participating centers. All participants provided written informed consent before enrollment.

### Participants and randomization

HCWs between 18 and 65 years of age without severe comorbidities, and no known history of SARS-CoV-2 infection (either laboratory-confirmed or self-reported) were eligible ^9^. All participants were primed with the Ad26.COV2.S vaccine 3 months before enrollment and were randomized in one of the following groups; Ad26.COV2.S/no boost, Ad26.COV2.S/Ad26.COV2.S, Ad26.COV2.S/mRNA-1273, or Ad26.COV2.S/BNT162b2. Participants were assigned to study groups in a 1:1:1:1 fashion; randomization was stratified by study site after obtaining written informed consent.

### Study design

Ad26.COV2.S-primed HCWs received a booster vaccination at study visit 1 (day 0) by injection in the deltoid muscle. The prime-boost interval was 84 days (−7 days / +21 days). Participants were blinded to the allocated vaccine by applying blinded etiquettes to conceal volume and appearance. The vaccines were administered according to the Summary of Product Characteristics. HCWs randomized to the Ad26.COV2.S/no boost group were informed of their allocation at study visit 1, and did not receive a placebo injection. Blood samples were collected at the timepoint of study visit 1 and 2 (0 and 28 days). Participants were unblinded for the booster vaccination by e-mail eight days after injection, after completing the reactogenicity questionnaires.

### Safety assessments

Safety assessments included monitoring of self-reported local and systemic reactions after the Ad26.COV2.S priming vaccination and different booster vaccinations by use of a modified 4-point US Food and Drug administration toxicity scale (0=no complaints, 1=mild complaints, does not interfere with daily activities, 2=moderate, interfere with daily activities or 3=severe, daily activities are no longer possible)^20^. Participants filled in electronic questionnaires retrospectively, ±84 days after the initial priming vaccination (all participants) and prospectively 7 days following the booster vaccination. Serious adverse events and solicited local or systemic reactions were self-reported (either through the questionnaire, by e-mail, or by phone). Safety monitoring (blood biochemistry and hematology assessment) was performed at days 0 and 28.

### Analyses of humoral and cellular immunity

The analysis of humoral and cellular immune responses is described in detail in the **Supplementary Material**. Briefly, SARS-CoV-2 nucleocapsid (N)-specific antibodies were measured to confirm that participants had not been previously exposed to SARS CoV-2. Spike (S)-specific binding antibodies were measured at 0 and 28 days after the booster vaccination using a quantitative anti-spike IgG assay (Liaison SARS-CoV-2 TrimericS IgG assay, DiaSorin, Italy) ^21,22^. Neutralizing capacity of antibodies against infectious SARS CoV-2 D614G was assessed in a plaque reduction neutralization test (PRNT) on Vero-E6 cells. SARS-CoV-2-specific T-cell responses were assessed by IFN-⍰ Release Assay (IGRA, QuantiFERON, Qiagen) at day 0 and 28 after booster vaccination as previously described ^23^.

### Statistical analysis

All statistical analyses are described in the **Supplementary Material**.

## Results

### Baseline characteristics

697 HCWs were screened for eligibility, of whom 461 were randomized for a booster vaccination; 27 individuals were excluded from further analyses **(Figure 1)**. All randomized HCWs included in the per protocol analysis (n=434) had their first study visit within the pre-defined period of 84 days (−7 and +21 days) and adhered to the timing between study visits **(Table 1)**. The final sample consisted of 280 (64.7%) women and 153 (35.3%) men with a median age of 40 years (IQR:30-50) and a median BMI of 23.9 (IQR 21.6-26.6). Baseline characteristics were not different between groups **(Table 1)**. At baseline, there were no significant differences with respect to S-specific IgG binding antibodies (p=0.39), neutralizing antibodies (p=0.55), and SARS-CoV-2-specific T-cell responses (p=0.92) across groups **(Figure 2A, 2C, 3A)**.

**Table 1.**
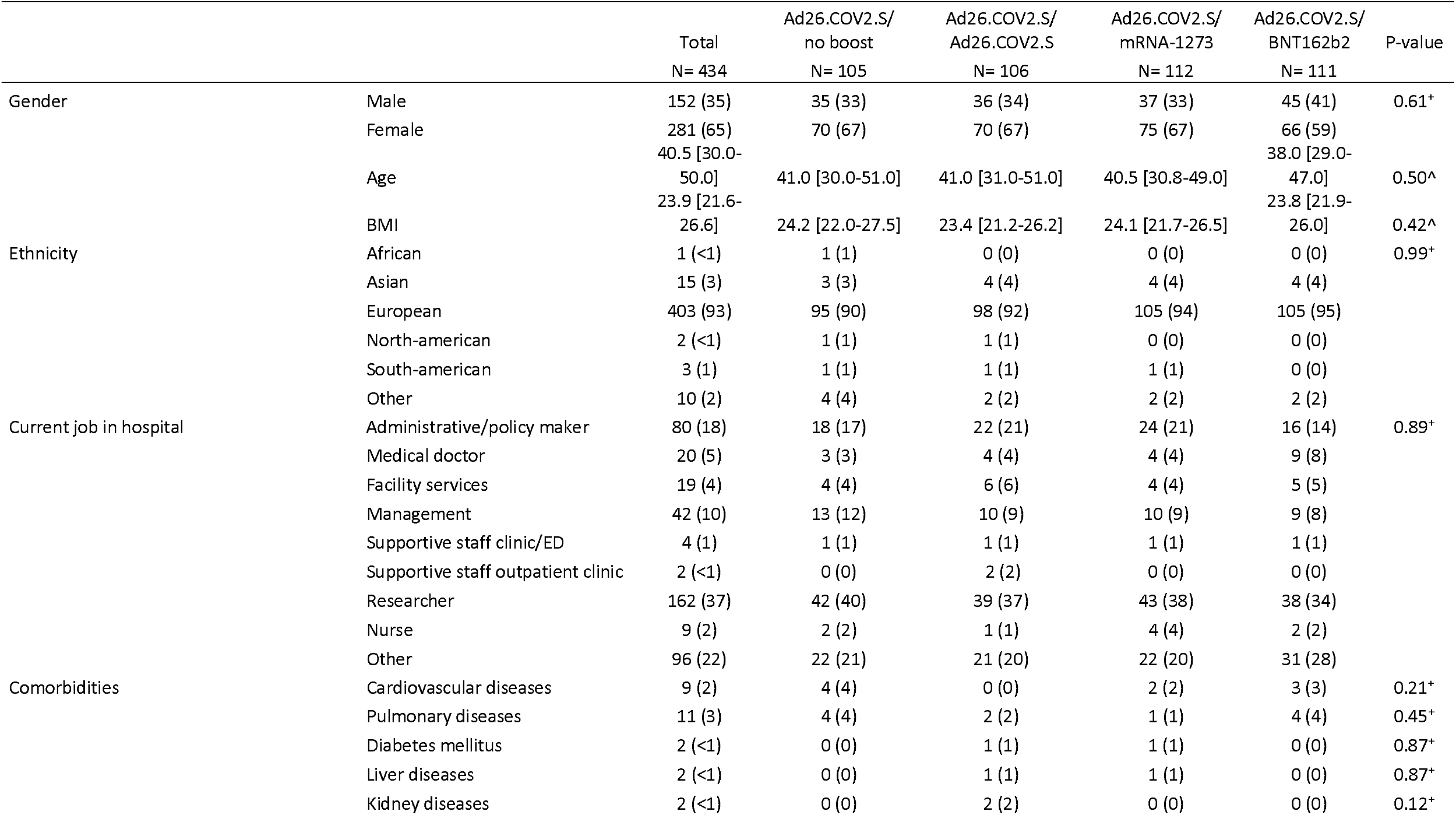
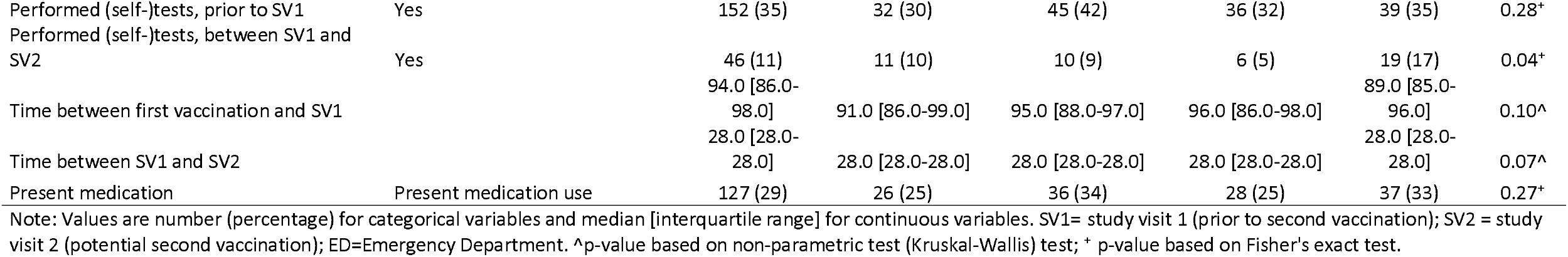
Baseline characteristics.

**Figure 1.**
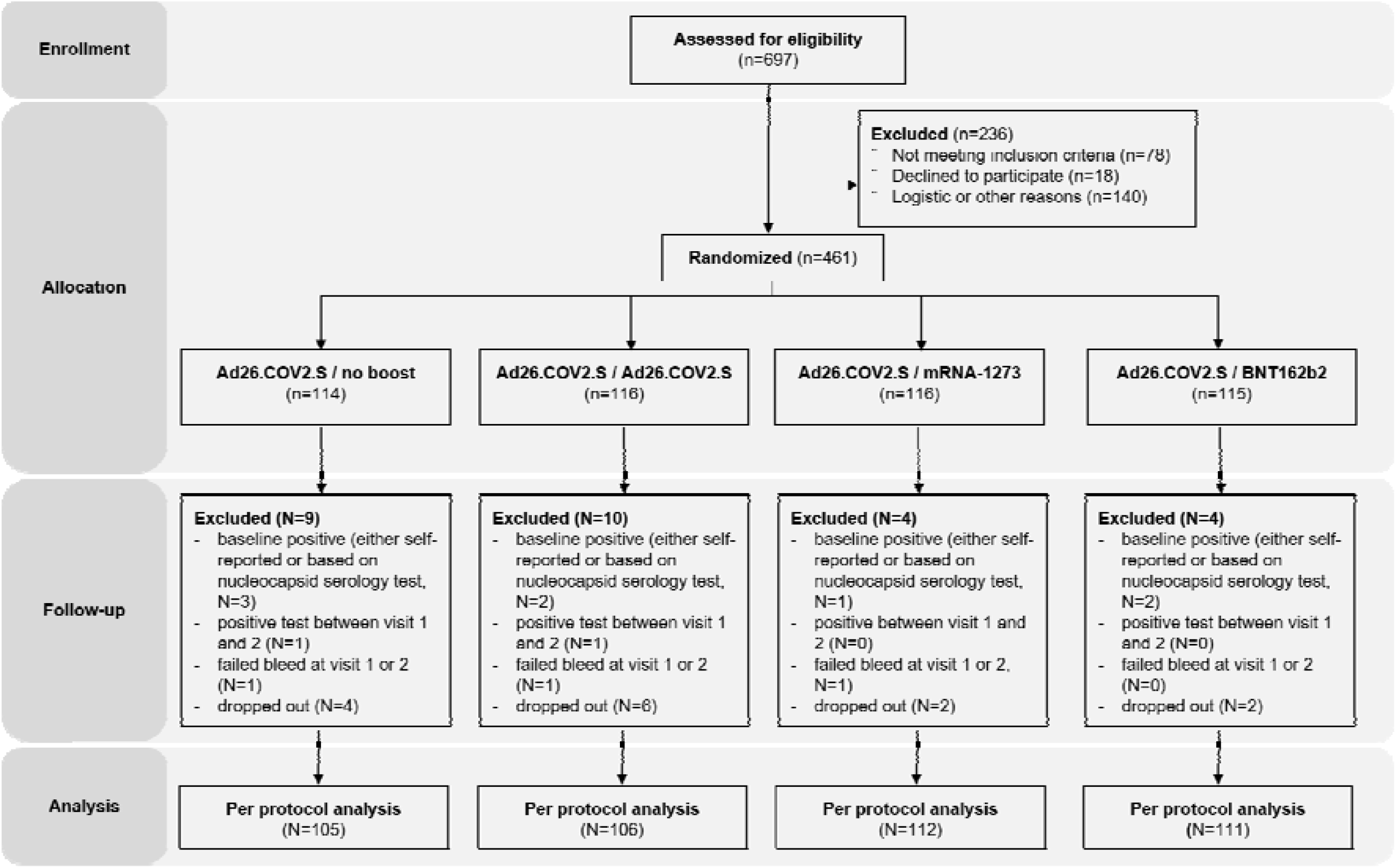
Study design and per protocol analysis. Flow diagram with the number of screened HCWs, randomization groups, follow-up, and inclusions in the per protocol analysis. After screening, a relatively high number of HCWs (N=140) dropped out for logistic or other reasons. This was mainly due to the strict schedule (84 days between prime and boost). After randomization, the number of HCWs who were lost-to-follow up were not statistically significant different across groups (p=0.19).

**Figure 2.**
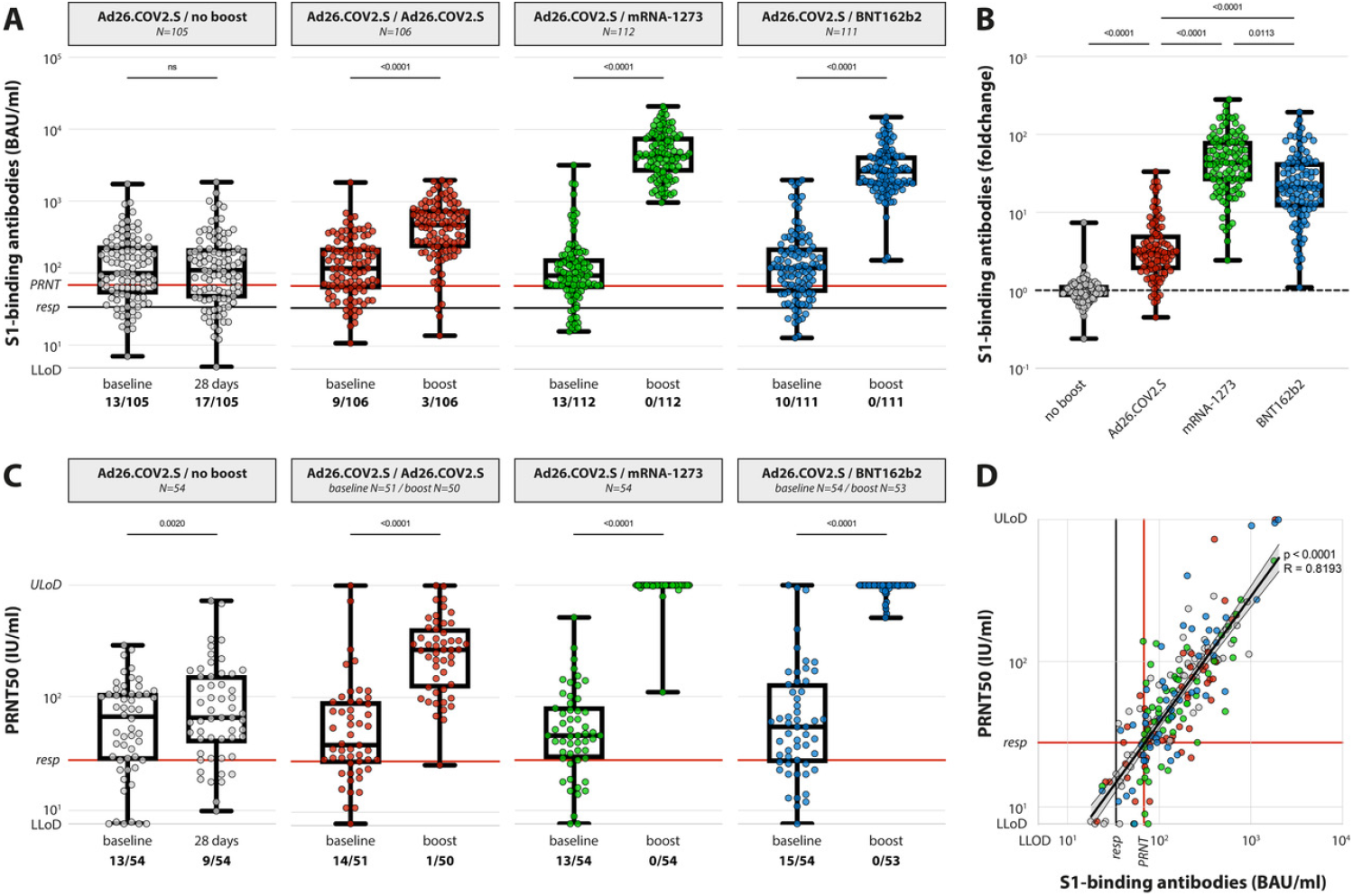
SARS-CoV-2-specific humoral responses. (A) Levels of binding S-specific antibodies at baseline and post booster vaccination in the 4 different groups. LLoD is 4.81 BAU/ml, responder [resp] cut-off is 33.8 BAU/ml (black line), levels >68.3 BAU/ml correlate to neutralizing capacity (red line, based on panel D). Data is presented as min-to-max box plots with individual values, timepoints were compared by Wilcoxon test. Number of participants below responder cut-off is indicated. (B) Individual fold changes were calculated by dividing the post-boost response by the pre-boost response. Data is presented as min-to-max box plots with individual values, groups were compared by Kruskal-Wallis test. (C) Levels of neutralizing antibodies at baseline and post booster vaccination in the 4 different groups. LLoD is 7.7 IU/ml, responder cut-off is set at 28.6 IU/ml. Data is presented as min-to-max box plots with individual values, timepoints were compared by Wilcoxon test. Number of participants below responder cut-off is indicated. (D) Correlation between binding S-specific antibodies and neutralizing antibodies. Spearman R = 0.83, p <0.001. Linear regression on log-transformed data was performed (Y=2.89*X-0.498), indicating that presence of neutralizing antibodies (>28.6) correlates with a binding antibody level >68.3 (red lines, resp).

### Rapid recall of S-specific antibodies in primed HCWs after booster vaccination

Levels of anti-S1 IgG binding antibodies were determined pre- and post-booster vaccination (days 0 and 28) by a quantitative assay **(Figure 2A, B)**. Based on a test cut-off for positivity at 33.8 BAU/ml (as per manufacturer’s instructions), 389 of 434 Ad26.COV2.S-primed HCWs had detectable S-binding antibodies at baseline (3 months after prime, 89.6% responders). We found no significant differences in percentage of participants without detectable binding antibodies across groups (p=0.75), with 12.4%, 8.4%, 11.6%, and 9% for no boost, Ad26.COV2.S boost, mRNA-1273 boost, and BNT162b2 boost, respectively.

Booster vaccinations led to a significant increase in S-specific binding antibodies in all groups compared to baseline levels (p<0.001 in all groups, **Figure 2A**). When an increase in antibody levels was calculated as a per participant fold change (dividing post-boost by pre-boost levels), boosting with either Ad26.COV2.S, mRNA-1273, or BNT162b2 led to a significant increase in antibodies compared to the Ad26.COV2.S single shot regimen (p<0.001 in all groups, **Figure 2B**). Notably, the heterologous mRNA-based booster vaccinations resulted in significantly higher binding antibody levels than homologous boost with Ad26.COV2.S (p<0.001), with mRNA-1273 performing slightly better than BTN162b2 (p=0.01). Booster vaccination with heterologous mRNA-based vaccines also led to a 100% response rate, whereas 3 of 106 participants in the Ad26.COV2.S boost group remained below the test cut-off for presence of binding antibodies.

### Neutralizing antibodies increase after homologous or heterologous booster vaccination

In addition to binding antibodies, SARS-CoV-2-specific neutralizing antibodies were measured by an infectious virus PRNT in participants from one academic center (Erasmus MC, N=213) **(Figure 2C, Supplementary Figure 1)**. Based on a test cut-off for positivity at 28.6 IU/ml (corresponding to a serum dilution of 1:40), 158 of 213 Ad26.COV2.S-primed HCWs had detectable neutralizing antibodies at baseline (3 months after prime, 74.2%); participants without detectable vaccine-induced neutralizing antibodies were equally divided over the study groups (24.1% no boost, 27.5% Ad26.COV2.S boost, 24.1% mRNA-1273 boost, and 27.8% BNT162b2 boost; p=0.95).

Booster vaccinations led to a significant increase in neutralizing antibodies in all groups compared to baseline levels (p<0.001 in all groups). After booster vaccination with any of the vaccines, neutralizing antibody levels in participants without detectable pre-booster neutralization were increased to above detection level with 1 exception in the Ad26.COV2.S boost group. Overall, heterologous mRNA-based booster vaccination increased neutralizing antibodies to higher levels when compared to Ad26.COV2.S booster vaccination (**Figure 2C, Supplementary Figure 1**).

### S-specific binding antibodies correlate to the presence of neutralizing antibodies

By performing a linear regression analysis on log-transformed binding and neutralizing antibody levels in pre-boost sera (Y=2.89*X-0.498), we were able to determine that the two are strongly correlated (Spearman R = 0.83, p <0.001) and that a binding antibody level of >68.3 BAU/ml correlates to the presence of neutralizing antibodies (>28.6 IU/ml) (**Figure 2D**). Re-analyzing the entire cohort for the presence of neutralizing antibodies with this arbitrary cut-off set at 68.3 BAU/ml indicated that 308 of 434 Ad26.COV2.S-primed HCW potentially had neutralizing antibodies at baseline, corresponding to 71.0%. Homologous booster vaccination increased neutralization response rate to 95.2%, whereas heterologous mRNA-based booster vaccination led to a 100% response rate regarding neutralizing antibodies.

### Rapid recall of T-cell responses in primed HCWs after booster vaccination

SARS-CoV-2-specific T-cell responses were measured pre- and post-booster vaccination (days 0 and 28) in whole blood at 3 participating centers in a random selection of samples (N=182) (**Figure 3A, B, Supplementary Figure 2**). Using a peptide pool covering the S protein (Ag2, Qiagen) and based on a test cut-off for positivity at 0.15 IU IFN-⍰/ml assessed by IGRA (per manufacturer’s instructions), 119 of 182 Ad26.COV2.S-primed HCWs had detectable T-cell responses at baseline.

**Figure 3.**
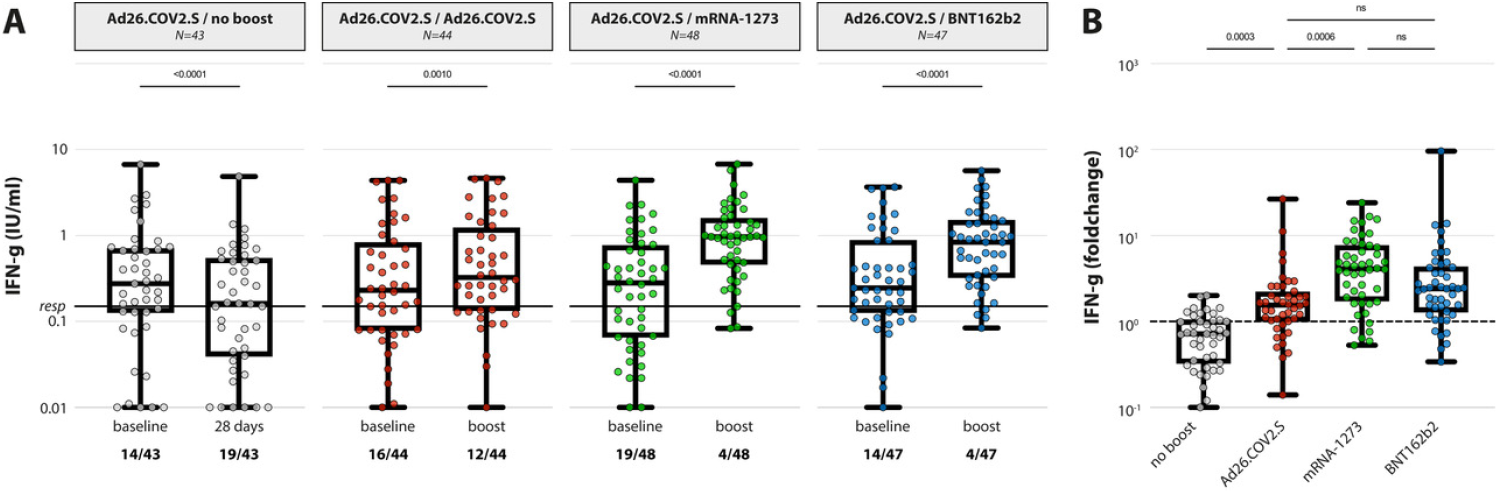
SARS-CoV-2-specific T-cell responses. (A) IFN-⍰ levels in plasma after stimulation of whole blood with an overlapping S pool (Ag2, Qiagen) at baseline and post booster vaccination in the 4 different groups. LLoD is 0.01 IU/ml, responder cut-off is 0.15 IU/ml. Data is presented as min-to-max box plots with individual values, timepoints were compared by Wilcoxon test. Number of participants below responder cut-off is indicated. (B) Individual fold changes were calculated by dividing the post-boost response by the pre-boost response. Data is presented as min-to-max box plots with individual values, groups were compared by Kruskal-Wallis test.

Booster vaccinations led to a rapid recall of T-cell responses in all groups compared to baseline levels (p=0.001 for Ad26.COV2.S boost, p<0.001 for mRNA-based boost) (**Figure 3A**). When an increase in T-cell responses was calculated as a per participant fold change, boosting with either Ad26.COV2.S, mRNA-1273, or BNT162b2 led to a significant increase in T-cells compared to the Ad26.COV2.S single shot regimen (p<0.001 in all groups, **Figure 3B**). Notably, heterologous booster vaccination with mRNA-1273 resulted in significantly higher T-cell responses than the homologous boost (p<0.001), but BNT162b2 booster vaccination did not. In terms of response rate, both mRNA-1273 and BNT162b2 boost (91.7% and 91.5% response rate, respectively) performed better than homologous Ad26.COV2.S boost (72.7%). Similar trends were observed with two other peptide pools (Ag1: receptor binding domain-specific T-cells, Ag3: S and other structural proteins, **Supplementary Figure 2**). SARS-CoV-2-specific T-cell responses significantly correlated to the presence of S-specific binding antibodies (**Supplementary Figure 3**).

### Reactogenicity

Reactogenicity data (severity and duration) retrospectively collected on the prime with Ad26.COV2.S is provided in **Supplementary Table 1 and 2**. Reactogenicity data (severity and duration) prospectively collected during seven days following the booster vaccination indicated increased severity of local (pain, redness, and swelling at injection site) and systemic (chills, fever, and muscle ache) reactions after heterologous booster vaccination with mRNA-1273 as compared to the other vaccination regimens (all p <0.01, **Figure 4**, actual data in **Supplementary Table 3 and 4**). The difference between regimens was most prominent on the day of boost and day 1 after vaccination (p<0.01). All adverse events were mild to moderate, did not require hospitalization, and symptoms generally disappeared within 48 hours, indicating that both homologous and heterologous regimens were well-tolerated.

**Figure 4.**
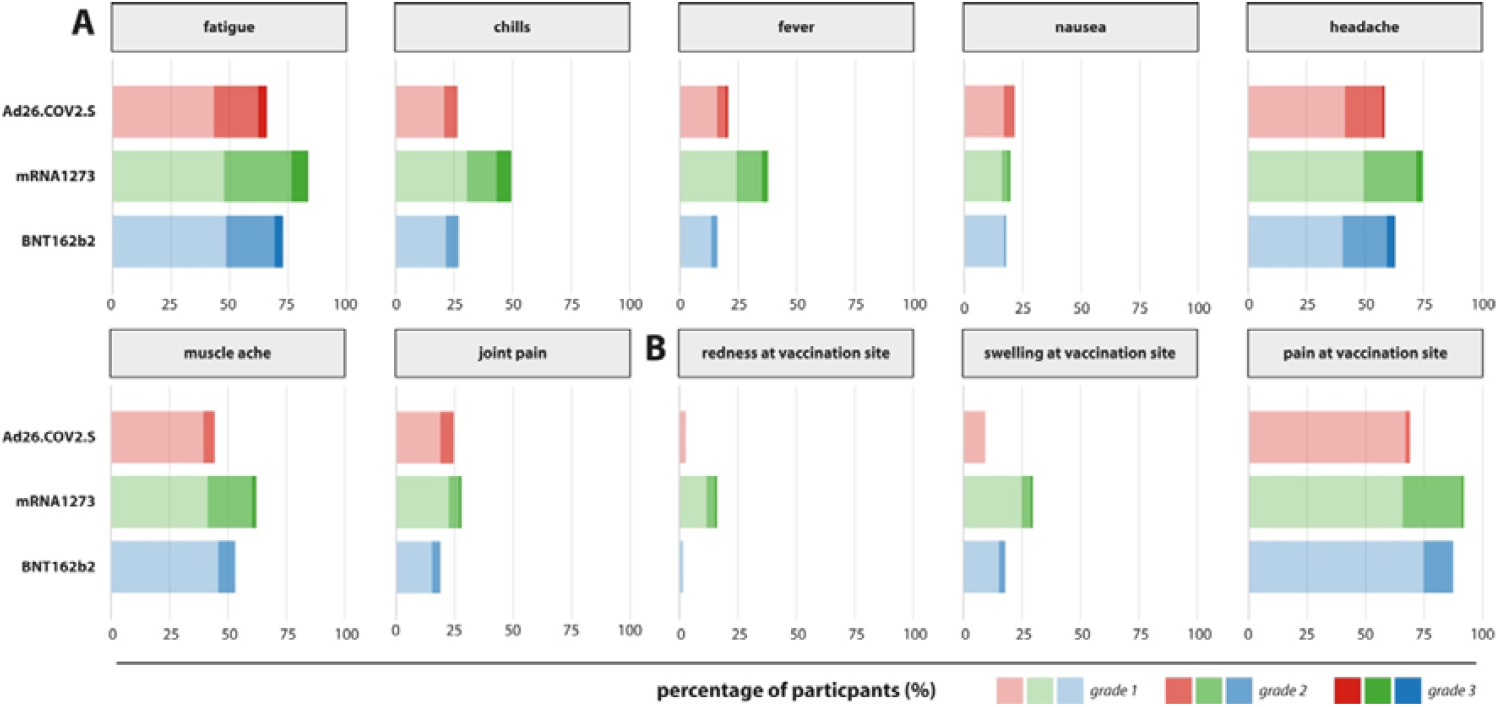
Systemic and local reactions after booster vaccination. Systemic (fatigue, chills, fever, nausea, headache, muscle ache, joint pain) and local (redness, swelling and / or pain at injection site) reactions were monitored in the seven days following boost. P-values for differences between the distribution of adverse events per group can be found in **Supplementary Table 4**.

## Discussion

In the SWITCH trial, we examined the immunogenicity and reactogenicity of homologous and heterologous booster vaccination regimens in previously Ad26.COV2.S-primed HCW, and compared these regimens to the approved Ad26.COV2.S single shot regimen. Both homologous and heterologous booster vaccinations led to an increase in SARS-CoV-2-specific binding antibodies, neutralizing antibodies and T-cell responses, but the increase was most prominent in heterologous regimens with mRNA-based COVID-19 vaccines mRNA1273 and BNT162b2.

Protection against a severe clinical course of COVID-19 with the currently approved Ad26.COV2.S single shot regimen is high ^3^, and it was recently reported that vaccination-induced immune responses can stably be detected up to 8 months post vaccination ^5^. However, in comparison to the mRNA-based vaccines approved in Europe, antibody responses induced by Ad26.COV2.S are lower than responses to mRNA-based vaccines, which raises the questions if booster vaccinations might be necessary to protect against currently and possibly future circulating SARS-CoV-2 variants of concern (VOC) that partially evade antibody responses such as the Beta and Delta variants ^7,8,24^.

A first aspect to evaluate is the safety of booster vaccinations. Heterologous ‘mixing and matching’ of approved vaccines has not been evaluated in phase 3 clinical trials, meaning that safety and reactogenicity of these vaccination regimens should be strictly evaluated in post-licensure studies. We did not observe any serious adverse events in this study; however, the sample size and study period is too small to observe the prevalence of rare adverse events. Potentially correlated to stronger boosting of immune responses, we found that mRNA1273-boosted participants perceived more mild local and systemic reactions. This suggests that mixing and matching Ad26.COV2.S with mRNA-based vaccines is safe and well-tolerated, as described by others ^18,19^.

The next step in decision making on the necessity of booster vaccinations is identifying a correlate of protection. Although well-defined correlates of protection against SARS CoV-2 infection have not yet been defined ^25^, neutralizing antibodies were shown to play an important role ^26^. A challenge is that assays to measure these responses are not standardized, making data from individual studies difficult to compare. Therefore, we propose an arbitrary cut-off of binding IgG antibodies that directly correlates to neutralizing capacity against D614G in Ad26.COV2.S-vaccinated individuals (68.3 BAU/ml), based on validation studies with an NIBSC reference standard and studies in COVID-19 patients ^27-29^. By doing so, we were able to combine the advantages of standardized high throughput analyses for binding antibodies with the biologically relevant measure of functional antibodies that inhibit binding of SARS-CoV-2 to host cells. By using this surrogate binding cut-off for neutralization, we determined that 71.0% of participants already had neutralizing antibodies against SARS-CoV-2 after a single Ad26.COV2.S injection. However, boosting significantly increased the levels. In this study, we measured neutralization of D614G, which is close to the virus used for development of the approved adenovirus- and mRNA-based vaccines, to specifically focus on vaccine responses. Based on published information on cross-neutralization of variants, higher neutralizing titers might be required to protect against (future) VOC.

In addition to antibody responses, induction and boosting of SARS CoV-2-specific T cells should be part of overall immunological assessments ^30^. Here, we show that SARS-CoV-2-specific T-cells were detectable in only 65.8% of Ad26.COV2.S-primed participants 3 months post vaccination. Especially boosting with mRNA-based vaccines led to a rapid increase in SARS-CoV-2-specific T-cells (100% response rate), indicating that immunity formed by the priming vaccination could rapidly be recalled by boosting. It was previously shown that SARS-CoV-2-specific T-cells equally recognize different variants of concern ^31^, and therefore especially the induction of T-cells is important in face of waning antibody levels and circulation of SARS-CoV-2 variants.

A potential limitation of our study is that we have exclusively evaluated booster vaccinations in young HCWs without severe comorbidities, and we may not be able translate these results to other populations. However, previous studies on homologous vaccination regimens show similar immunogenicity data comparing younger (18–55 years) and older (>55 years) adults ^32-34^. Furthermore, we evaluated the heterologous booster vaccination regimens in a three-month interval from the priming vaccination, to allow a comparison with ChAdOx1 nCoV-19 trials ^35^, but the optimal prime-boost interval remains to be investigated. The homologous prime-boost intervals in the Ad26.COV2.S trials published after inception of the SWITCH trial varied from 2 to 6 months ^6,36^, and suggest that late boosting might be more effective. This would further enhance the effect observed in our study.

In conclusion, single dose Ad26.COV2.S vaccination adequately primes the immune system. We now show that in face of possible waning immunity and circulation of SARS-CoV-2 variants, these responses can be boosted most efficiently with mRNA vaccines. Upon boosting, an increased VE against infection and transmission is likely, but future studies will need to show the added value of boosting on VE against severe disease. Discussion on the need for booster vaccination should also consider timing, the target population, the level of SARS CoV-2 circulation, and the global inequity in vaccine access.

## Supporting information

Consort statement

## Data Availability

All data produced in the present study are available upon reasonable request to the authors

## On behalf of the SWITCH research group

**Erasmus MC**

N. Tjon

K. van Grafhorst

L.P.M. van Leeuwen

S. Bogers

F. de Wilt

L. Gommers

S. Scherbeijn

A.C.P. Lamoré

**Amsterdam UMC**

A.M. Harskamp

I. Maurer

A.F. Girigorie

B. D. Boeser-Nunnink

M.M. Mangas Ruiz

**UMCG**

R. Akkerman

M. Beukema

J.J. de Vries-Idema

J. Zuidema

**LUMC**

J.A. Vlot

P.H. Verbeek -Menken

## References

1. Polack FP, Thomas SJ, Kitchin N, et al. Safety and Efficacy of the BNT162b2 mRNA Covid-19 Vaccine. N Engl J Med 2020;383:2603–15.

2. Baden LR, El Sahly HM, Essink B, et al. Efficacy and Safety of the mRNA-1273 SARS-CoV-2 Vaccine. N Engl J Med 2021;384:403–16.

3. Sadoff J, Gray G, Vandebosch A, et al. Safety and Efficacy of Single-Dose Ad26.COV2.S Vaccine against Covid-19. N Engl J Med 2021.

4. Voysey M, Costa Clemens SA, Madhi SA, et al. Safety and efficacy of the ChAdOx1 nCoV-19 vaccine (AZD1222) against SARS-CoV-2: an interim analysis of four randomised controlled trials in Brazil, South Africa, and the UK. Lancet 2021: 99–111.

5. Barouch DH, Stephenson KE, Sadoff J, et al. Durable humoral and cellular immune responses 8 months after Ad26. COV2. S vaccination. N Engl J Med 2021;385:951–3.

6. Sadoff J, Le Gars M, Cardenas V, et al. Durability of antibody responses elicited by a single dose of Ad26.COV2.S and substantial increase following late boosting. medRxiv 2021:2021.08.25.21262569.

7. van Gils MJ, Lavell AHA, van der Straten K, et al. Four SARS-CoV-2 vaccines induce quantitatively different antibody responses against SARS-CoV-2 variants. medRxiv 2021:2021.09.27.21264163.

8. Collier AY, Yu J, McMahan K, et al. Differential Kinetics of Immune Responses Elicited by Covid-19 Vaccines. N Engl J Med 2021.

9. Sablerolles RSG, Goorhuis A, GeurtsvanKessel C, et al. Heterologous Ad26.COV2.S Prime and mRNA-Based Boost COVID-19 Vaccination Regimens: The SWITCH Trial Protocol. Front Immunol 2021.

10. Spencer AJ, McKay PF, Belij-Rammerstorfer S, et al. Heterologous vaccination regimens with self-amplifying RNA and adenoviral COVID vaccines induce robust immune responses in mice. Nat Commun 2021;12:2893.

11. He Q, Mao Q, An C, et al. Heterologous prime-boost: breaking the protective immune response bottleneck of COVID-19 vaccine candidates. Emerging Microbes & Infections 2021;10:629–37.

12. Duarte-Salles T, Prieto-Alhambra D. Heterologous vaccine regimens against COVID-19. The Lancet 2021;398:94–5.

13. Hacisuleyman E, Hale C, Saito Y, et al. Vaccine breakthrough infections with SARS-CoV-2 variants. N Engl J Med 2021;384:2212–8.

14. Krause PR, Fleming TR, Longini IM, et al. SARS-CoV-2 Variants and Vaccines. N Engl J Med 2021.

15. Liu X, Shaw RH, Stuart ASV, et al. Safety and immunogenicity of heterologous versus homologous prime-boost schedules with an adenoviral vectored and mRNA COVID-19 vaccine (Com-COV): a single-blind, randomised, non-inferiority trial. The Lancet 2021;398:856–69.

16. Hillus D, Schwarz T, Tober-Lau P, et al. Safety, reactogenicity, and immunogenicity of homologous and heterologous prime-boost immunisation with ChAdOx1 nCoV-19 and BNT162b2: a prospective cohort study Lancet Respir Med 2021.

17. Borobia AM, Carcas AJ, Pérez-Olmeda M, et al. Immunogenicity and reactogenicity of BNT162b2 booster in ChAdOx1-S-primed participants (CombiVacS): a multicentre, open-label, randomised, controlled, phase 2 trial. The Lancet 2021;398:121–30.

18. Shaw RH, Stuart A, Greenland M, Liu X, Van-Tam JSN, Snape MD. Heterologous prime-boost COVID-19 vaccination: initial reactogenicity data. The Lancet 2021;397:2043–6.

19. Atmar RL, Lyke KE, Deming ME, et al. Heterologous SARS-CoV-2 Booster Vaccinations: Preliminary Report. medRxiv 2021:2021.10.10.21264827.

20. Food and Drug Administration (FDA). Toxicity grading scale for healthy adults and adolescent volunteers enrolled in preventive vaccine clinical trials. Available from: https://www.fda.gov/media/73679/download [accessed 1st April 2021].

21. Mahmoud SA, Ganesan S, Bissar S, Zaher W. Evaluation of serological tests for detecting SARS-CoV-2 antibodies: implementation in assessing post vaccination status. medRxiv 2021.

22. Leuzinger K, Osthoff M, Dräger S, et al. Comparing immunoassays for SARS-Coronavirus-2 antibody detection in patients with and without laboratory-confirmed SARS-Coronavirus-2 infection. J Clin Microbiol 2021:JCM0138121.

23. Sanders J-S, Bemelman FJ, Messchendorp AL, et al. The RECOVAC Immune-Response Study The immunogenicity, tolerability and safety of COVID-19 vaccination in patients with chronic kidney disease, on dialysis, or living with a kidney transplant. Transplantation 2021;forthcoming.

24. Rishi RG, Mark MP, Sokratis AA, et al. mRNA vaccines induce durable immune memory to SARS-CoV-2 and variants of concern. Science;0:eabm0829.

25. Krammer F. A correlate of protection for SARS-CoV-2 vaccines is urgently needed. Nature medicine 2021;27:1147–8.

26. Khoury DS, Cromer D, Reynaldi A, et al. Neutralizing antibody levels are highly predictive of immune protection from symptomatic SARS-CoV-2 infection. Nature medicine 2021:1–7.

27. van Kampen JJA, van de Vijver D, Fraaij PLA, et al. Duration and key determinants of infectious virus shedding in hospitalized patients with coronavirus disease-2019 (COVID-19). Nat Commun 2021;12:267.

28. Okba NMA, Muller MA, Li W, et al. Severe Acute Respiratory Syndrome Coronavirus 2-Specific Antibody Responses in Coronavirus Disease Patients. Emerg Infect Dis 2020;26:1478–88.

29. GeurtsvanKessel CH, Okba NMA, Igloi Z, et al. An evaluation of COVID-19 serological assays informs future diagnostics and exposure assessment. Nat Commun 2020;11:3436.

30. Tan AT, Lim JME, Le Bert N, et al. Rapid measurement of SARS-CoV-2 spike T cells in whole blood from vaccinated and naturally infected individuals. The Journal of Clinical Investigation 2021;131.

31. Geers D, Shamier MC, Bogers S, et al. SARS-CoV-2 variants of concern partially escape humoral but not T-cell responses in COVID-19 convalescent donors and vaccinees. Science Immunology 2021.

32. Anderson EJ, Rouphael NG, Widge AT, et al. Safety and Immunogenicity of SARS-CoV-2 mRNA-1273 Vaccine in Older Adults. N Engl J Med 2020;383:2427–38.

33. Walsh EE, Frenck Jr RW, Falsey AR, et al. Safety and immunogenicity of two RNA-based Covid-19 vaccine candidates. N Engl J Med 2020;383:2439–50.

34. Ramasamy MN, Minassian AM, Ewer KJ, et al. Safety and immunogenicity of ChAdOx1 nCoV-19 vaccine administered in a prime-boost regimen in young and old adults (COV002): a single-blind, randomised, controlled, phase 2/3 trial. The Lancet 2020:1979–93.

35. Voysey M, Clemens SAC, Madhi SA, et al. Single-dose administration and the influence of the timing of the booster dose on immunogenicity and efficacy of ChAdOx1 nCoV-19 (AZD1222) vaccine: a pooled analysis of four randomised trials. The Lancet 2021;397:881–91.

36. Stephenson KE, Le Gars M, Sadoff J, et al. Immunogenicity of the Ad26.COV2.S Vaccine for COVID-19. JAMA 2021;325:1535–44.

