## Supplementary Material for "Immunogenicity and reactogenicity of booster vaccinations after Ad26.COV2.S priming"

**Supplementary Methods**

**Serology**

Nucleocapsid-specific antibodies were measured on an Abbott Architect instrument using the Abbott SARS-CoV-2 IgG assay following the manufacturer’s instructions ^1^. Qualitative results and index values reported by the instrument were used in analyses, using the manufacturer’s cut-off for positivity, i.e., 1.4 S/CO. Antibodies against the SARS CoV-2 Spike (S) protein were measured by Liaison SARS CoV-2 TrimericS IgG assay (DiaSorin, Italy), with a lower limit of detection of 4.81 BAU/ml and a cut-off for positivity at 33.8 BAU/ml. The assay was performed following the manufacturer’s instructions.

**Plaque reduction neutralization test (PRNT_50_)**

PRNT was previously described ^2,3^. SARS CoV-2 D614G used in the assay was isolated from a diagnostic specimen at the Department of Viroscience, Erasmus MC, cultured, and subsequently sequenced to rule out additional mutations in the S protein: D614G [Global Initiative on Sharing All Influenza Data (GISAID): hCov-19/Netherlands/ZH-EMC-2498]. Heat-inactivated sera were two-fold diluted in Dulbecco’s modified Eagle’s medium (DMEM) supplemented with NaHCO3, Hepes buffer, penicillin, streptomycin, and 1% FBS, starting at a dilution of 1:10 in 60ul. 60ul of virus suspension was added to each well and incubated at 37°C for 1 hour. After 1 hour of incubation, the virus-antibody mixtures were transferred onto Vero-E6 cells and incubated for 8 hours. After incubation, cells were fixated in 10% formaldehyde and plaques were stained with polyclonal rabbit anti–SARS-CoV-2 nucleocapsid antibody (Sino Biological) and a secondary peroxidase-labeled goat anti-rabbit IgG (Dako). Signal was developed by using a precipitate-forming 3,3′,5,5′-tetramethylbenzidine substrate (TrueBlue; Kirkegaard & Perry Laboratories) and the number of infected cells was counted per well by using an ImmunoSpot Image Analyzer (CTL Europe GmbH). The dilution that would yield 50% reduction of plaques (PRNT_50_) compared with the infection control (included on all plates) was estimated by determining the proportionate distance between two dilutions from which an end point titer was calculated. Infection controls and positive serum controls were included on each plate, the NIBSC standard was included in two separate experiments.

All neutralization titers were expressed relative to the NIBSC standard in IU/ml. The lower limit of detection of the assay was 7.7 IU/ml (corresponding to a serum dilution of 1:10), but based on assay validation a cut off of 28.6 IU/ml (corresponding to a serum dilution of 1:40) was considered as a minimal vaccine response. The upper limit of detection was 984 IU/ml (corresponding to a serum dilution of 1:2560). All pre-boost data within the dynamic range of the assay were used to correlate binding IgG levels to neutralizing antibodies (**Figure 2D**). Based on a minimal neutralizing titer of 28.6 IU/ml, a threshold of 68.3 BAU/mL was defined for binding anti-S IgG that correlates to neutralizing capacity (**Figure 2D**).

**INF-ɣ Release Assay**

The SARS-CoV-2-specific T cell response was measured by commercially available IFN-ɣ Release Assay (IGRA, QuantiFERON, Qiagen) in whole blood as previously described ^4^. In short, heparinized whole blood obtained at day 0 and at 28 days after the booster vaccination was incubated with three different SARS-CoV-2 antigens for 20-24h using a combination of peptides stimulating both CD4^+^ and CD8^+^ T-cells (Ag1, Ag2, Ag3, QuantiFERON, Qiagen). After incubation, plasma was obtained and IFN-ɣ production in response to the antigens was measured by ELISA. Results are expressed in IU IFN-ɣ/ml after subtraction of the NIL control values as interpolated from a standard calibration curve. Lower limit of detection in this assay is set at 0.01 IU/ml, responder cut-off is 0.15 IU/ml (per manufacturer’s instructions).

**Statistical analysis**

The power calculation for analysis of the primary endpoint was based on available data from the Com-COV1 and CombiVacs studies ^5,6^ as described in our protocol (per protocol analysis, with list wise deletion) ^7^. Continuous variables are presented as median (interquartile range). Comparing differences between multiple groups was done by a Kruskal-Wallis test, whereas comparing two groups was done using a non-parametric Wilcoxon rank test. Categorical variables are presented as numbers or percentages. Comparing frequencies between groups was done using a Fisher’s exact test. The primary (SARS-CoV-2-specific binding antibodies at day 0 and 28 days after the boost) and secondary end points were presented as min-max boxplots per group including all individual datapoints. Additionally, we calculated the per donor antibody and T-cell fold changes by dividing values post boost by values pre boost for group comparisons. Finally, we compared the reactogenicity endpoints per group with similar statistical comparisons as performed for baseline characteristics. Spearman correlation was calculated between binding and neutralizing antibodies, and between binding antibodies and IGRA. Linear regression on log-transformed data was performed. Statistical analyses were performed using GraphPad PRISM, version 9.1.2 and R studio, version 4.0.5 (R foundation for statistical computing).

**Supplementary Figures**

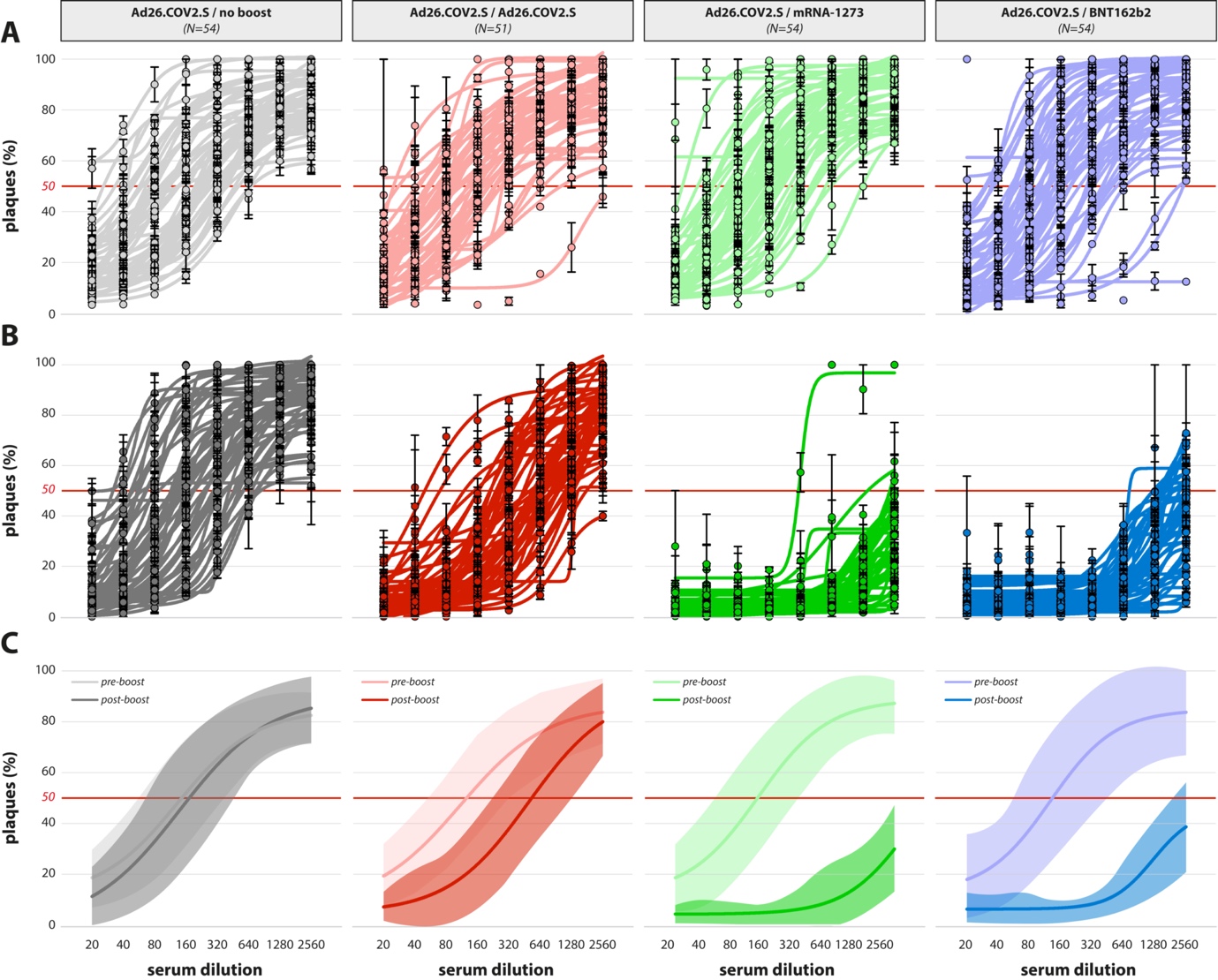

**Supplementary Figure 1. Normalized S-curves for determining PRNT_50_.** (A, B) Sera were 2-fold diluted starting at a dilution of 1:10 in duplicate, followed by adding virus suspension. Percentage plaque reduction compared to the infection control was calculated per dilution and plotted as log (inhibitor) vs response curve with four parameter variable slope using GraphPad Prism 9.02. The dilution that would yield 50% reduction of plaques compared to the infection control was estimated by determining the proportionate distance between two dilutions, from which an endpoint titer was calculated. (A) Normalized S-curves pre-boost and (B) normalized S-curves post-boost per study group. (C) Mean plaque reduction values per timepoint and per study group ± SD in shown as shaded area. Light color coding reflects PRNT pre-boost, dark color coding reflects PRNT post-boost.

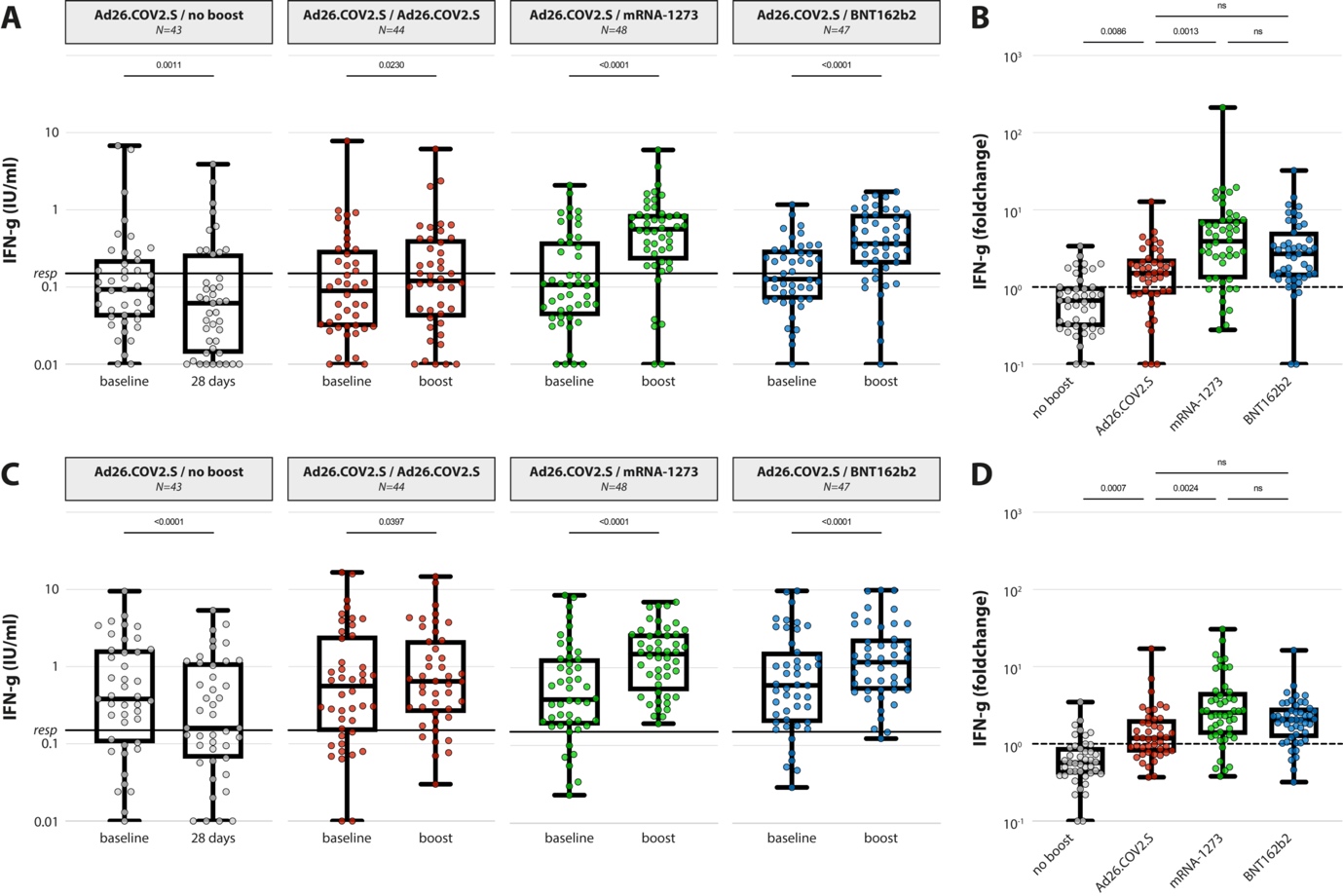

**Supplementary Figure 2. SARS-CoV-2-specific T-cell responses.** (A, C) IFN-ɣ levels in plasma after stimulation of whole blood with an overlapping S pool (panel A shows Ag1 or RBD-specific T-cell responses, panel C shows Ag3 or multiple protein specific T-cell responses) at baseline and post booster vaccination in the 4 different groups. LLoD is 0.01 IU/ml, responder cut-off is 0.15 IU/ml. Data is presented as min-to-max box plots with individual values, timepoints were compared by Wilcoxon test. (B, D) Individual fold changes were calculated by dividing the post-boost response by the pre-boost response (panel B shows Ag1, panel D shows Ag3, Qiagen). Data is presented as min-to-max box plots with individual values, groups were compared by Kruskal-Wallis.

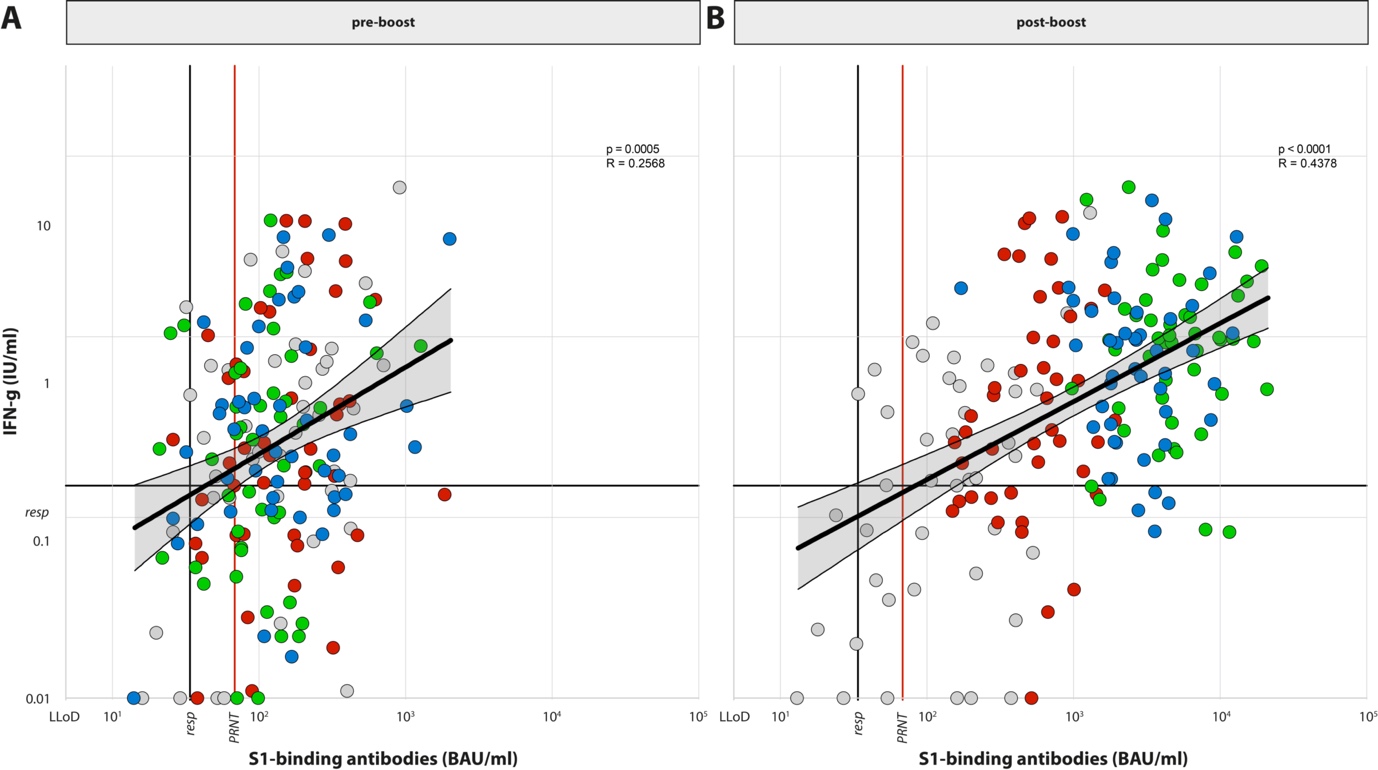

**Supplementary Figure 3. Correlation between binding antibodies and IGRA.** Correlation between binding S-specific antibodies and IGRA (pre- and post-boost (A, B). Spearman R was calculated Linear regression on log-transformed data was performed. Color coding is similar throughout figures, and represents the different study groups (grey = no boost, red = Ad26.COV2.S boost, green = mRNA-1273 boost, blue = BNT162b2 boost).

**Supplementary Tables**

**Supplementary table 1. Severity of the side-effects after Ad26.COV2.S-prime (retrospectively collected).**

| In the seven days after your second vaccination did you suffer from one of the following side-effects: | | Ad26.COV2.S/ no boost | Ad26.COV2.S/ Ad26.COV2.S | Ad26.COV2.S/ mRNA-1273 | Ad26.COV2.S/ BNT162b2 |  |
| --- | --- | --- | --- | --- | --- | --- |
|  |  | N=105 | N= 106 | N= 112 | N= 111 | p-value |
| Fatigue | Not at all | 27 (26) | 25 (24) | 40 (36) | 33 (30) | 0.65 |
|  | A little, but did not hinder my daily activity | 49 (47) | 42 (40) | 39 (35) | 44 (40) |  |
|  | Quite, it hindered my daily activity | 24 (23) | 34 (32) | 28 (25) | 28 (25) |  |
|  | Significantly, I could not perform my daily activity | 5 (5) | 5 (5) | 5 (4) | 6 (5) |  |
| Chills | Not at all | 53 (50) | 69 (65) | 65 (58) | 64 (58) | 0.25 |
|  | A little, but did not hinder my daily activity | 33 (31) | 18 (17) | 19 (17) | 21 (19) |  |
|  | Quite, it hindered my daily activity | 13 (12) | 15 (14) | 19 (17) | 18 (16) |  |
|  | Significantly, I could not perform my daily activity | 6 (6) | 4 (4) | 9 (8) | 8 (7) |  |
| Fever | Not at all | 68 (65) | 71 (67) | 71 (63) | 70 (63) | 0.97 |
|  | 38.0-38.4 | 21 (20) | 24 (23) | 26 (23) | 27 (24) |  |
|  | 38.5-38.9 | 10 (10) | 7 (7) | 12 (11) | 9 (8) |  |
|  | >39.0 | 6 (6) | 4 (4) | 3 (3) | 5 (5) |  |
| Nausea | Not at all | 92 (88) | 91 (86) | 96 (86) | 96 (86) | 0.77 |
|  | A little, but did not hinder my daily activity | 10 (10) | 10 (9) | 12 (11) | 10 (9) |  |
|  | Quite, it hindered my daily activity | 3 (3) | 5 (5) | 3 (3) | 2 (2) |  |
|  | Significantly, I could not perform my daily activity | 0 ()) | 0 (0) | 1 (1) | 3 (3) |  |
| Headache | Not at all | 50 (48) | 58 (55) | 51 (46) | 54 (49) | 0.78 |
|  | A little, but did not hinder my daily activity | 38 (36) | 33 (31) | 39 (35) | 34 (31) |  |
|  | Quite, it hindered my daily activity | 12 (11) | 12 (11) | 19 (17) | 15 (14) |  |
|  | Significantly, I could not perform my daily activity | 5 (5) | 3 (3) | 3 (3) | 8 (7) |  |
| Muscle ache | Not at all | 46 (44) | 49 (46) | 57 (51) | 62 (56) | 0.63 |
|  | A little, but did not hinder my daily activity | 43 (41) | 42 (40) | 40 (36) | 32 (29) |  |
|  | Quite, it hindered my daily activity | 14 (13) | 11 (10) | 14 (13) | 14 (13) |  |
|  | Significantly, I could not perform my daily activity | 2 (2) | 4 (4) | 1 (1) | 3 (3) |  |
| Joint pain | Not at all | 78 (74) | 82 (77) | 84 (75) | 89 (80) | 0.67 |
|  | A little, but did not hinder my daily activity | 21 (20) | 17 (16) | 23 (21) | 13 (12) |  |
|  | Quite, it hindered my daily activity | 5 (5) | 5 (2) | 5 (4) | 8 (7) |  |
|  | Significantly, I could not perform my daily activity | 1 (1) | 2 (2) | 0 (0) | 1 (1) |  |
| Red spot at vaccination site | Not at all | 96 (91) | 98 (92) | 107 (96) | 108 (97) | 0.28 |
|  | 2.5-5.0 cm | 8 (7) | 8 (8) | 5 (4) | 3 (3) |  |
|  | 5.1-10.0 cm | 1 (1) | 0 (0) | 0 (0) | 0 (0) |  |
|  | >10.0 cm | 0 (0) | 0 (0) | 0 (0) | 0 (0) |  |
| Swelling at vaccination site | Not at all | 95 (90) | 99 (93) | 102 (91) | 105 (95) | 0.63 |
|  | 2.5-5.0 cm | 10 (10) | 7 (7) | 10 (9) | 6 (5) |  |
|  | 5.1-10.0 cm | 0 (0) | 0 (0) | 0 (0) | 0 (0) |  |
|  | >10.0 cm | 0 (0) | 0 (0) | 0 (0) | 0 (0) |  |
| Pain at vaccination site | Not at all | 38 (26) | 35 (33) | 42 (38) | 30 (27) | 0.56 |
|  | A little, but did not hinder my daily activity | 66 (63) | 68 (64) | 68 (61) | 78 (70) |  |
|  | Quite, it hindered my daily activity | 1 (1) | 3 (3) | 1 (1) | 3 (0) |  |
|  | Significantly, I could not perform my daily activity | 0 (0) | 0 (0) | 1 (1) | 0 (0) |  |
| Note: Values are number (percentage). P-values are based on Fisher’s exact test. Bold p-values are significant at the 1% alpha level. | | | | | | |

**Supplementary table 2. Timing of side-effects over seven days following Ad26.COV2.S-prime (retrospectively collected).**

| In the seven days after your first vaccination did you suffer from one of the following side-effects: | | Ad26.COV2.S/no boost | Ad26.COV2.S/ Ad26.COV2.S | Ad26.COV2.S/ mRNA-1273 | Ad26.COV2.S/ BNT162b2 | p-value |
| --- | --- | --- | --- | --- | --- | --- |
|  |  | N= 105 | N= 106 | N= 112 | N= 111 |  |
| Fatigue | Day of vaccination | 28 (27) | 27 (25) | 30 (27) | 36 (32) | 0.67 |
|  | Day 1 | 70 (67) | 66 (62) | 63 (56) | 64 (58) | 0.38 |
|  | Day 2 | 35 (33) | 27 (25) | 30 (27) | 35 (32) | 0.54 |
|  | Day 3 | 9 (9) | 9 (8) | 9 (8) | 11 (10) | 0.97 |
|  | Day 4 | 8 (8) | 6 (6) | 5 (4) | 5 (5) | 0.73 |
|  | Day 5 | 4 (4) | 6 (6) | 1 (1) | 3 (3) | 0.22 |
|  | Day 6 | 2 (2) | 5 (5) | 1 (1) | 2 (2) | 0.35 |
|  | Day 7 | 1 (1) | 5 (5) | 1 (1) | 2 (2) | 0.26 |
| Chills | Day of vaccination | 36 (34) | 19 (18) | 36 (32) | 33 (30) | 0.03 |
|  | Day 1 | 36 (34) | 24 (23) | 20 (18) | 25 (23) | 0.04 |
|  | Day 2 | 4 (4) | 0 (0) | 0 (0) | 5 (5) | **0.01** |
|  | Day 3 | 2 (2) | 1 (1) | 1 (1) | 0 (0) | 0.43 |
|  | Day 4 | 0 (0) | 1 (1) | 1 (1) | 0 (0) | 0.87 |
|  | Day 5 | 0 (0) | 0 (0) | 0 (0) | 0 (0) | NA |
|  | Day 6 | 0 (0) | 0 (0) | 0 (0) | 0 (0) | NA |
|  | Day 7 | 0 (0) | 0 (0) | 0 (0) | 0 (0) | NA |
| Fever | Day of vaccination | 26 (25) | 19 (18) | 26 (23) | 25 (23) | 0.66 |
|  | Day 1 | 26 (25) | 24 (23) | 25 (22) | 31 (28) | 0.76 |
|  | Day 2 | 3 (3) | 2 (2) | 3 (3) | 5 (5) | 0.72 |
|  | Day 3 | 1 (1) | 0 (0) | 1 (1) | 0 (0) | 0.62 |
|  | Day 4 | 0 (0) | 0 (0) | 1 (1) | 0 (0) | 1.00 |
|  | Day 5 | 0 (0) | 0 (0) | 0 (0) | 0 (0) | NA |
|  | Day 6 | 0 (0) | 0 (0) | 0 (0) | 0 (0) | NA |
|  | Day 7 | 0 (0) | 0 (0) | 0 (0) | 0 (0) | NA |
| Nausea | Day of vaccination | 5 (5) | 7 (7) | 8 (7) | 9 (8) | 0.80 |
|  | Day 1 | 9 (9) | 9 (8) | 10 (9) | 9 (8) | 1.00 |
|  | Day 2 | 2 (2) | 5 (5) | 1 (1) | 4 (4) | 0.30 |
|  | Day 3 | 1 (1) | 2 (2) | 1 (1) | 1 (1) | 0.88 |
|  | Day 4 | 1 (1) | 0 (0) | 1 (1) | 2 (2) | 0.76 |
|  | Day 5 | 1 (1) | 0 (0) | 0 (0) | 0 (0) | 0.24 |
|  | Day 6 | 1 (1) | 0 (0) | 0 (0) | 0 (0) | 0.24 |
|  | Day 7 | 1 (1) | 0 (0) | 0 (0) | 0 (0) | 0.24 |
| Headache | Day of vaccination | 19 (18) | 18 (17) | 25 (22) | 28 (25) | 0.42 |
|  | Day 1 | 47 (45) | 36 (34) | 46 (41) | 48 (43) | 0.39 |
|  | Day 2 | 16 (15) | 19 (18) | 11 (10) | 15 (14) | 0.37 |
|  | Day 3 | 3 (3) | 7 (7) | 3 (3) | 3 (3) | 0.42 |
|  | Day 4 | 1 (1) | 3 (3) | 3 (3) | 1 (1) | 0.63 |
|  | Day 5 | 0 (0) | 3 (3) | 1 (1) | 0 (0) | 0.10 |
|  | Day 6 | 0 (0) | 1 (1) | 0 (0) | 0 (0) | 0.49 |
|  | Day 7 | 0 (0) | 2 (2) | 0 (0) | 0 (0) | 0.12 |
| Muscle ache | Day of vaccination | 24 (23) | 14 (13) | 29 (26) | 18 (16) | 0.07 |
|  | Day 1 | 55 (52) | 49 (46) | 45 (40) | 42 (38) | 0.14 |
|  | Day 2 | 25 (24) | 29 (27) | 17 (15) | 21 (19) | 0.13 |
|  | Day 3 | 12 (11) | 11 (10) | 8 (7) | 8 (7) | 0.59 |
|  | Day 4 | 5 (5) | 6 (6) | 7 (6) | 1 (1) | 0.14 |
|  | Day 5 | 2 (2) | 3 (3) | 4 (4) | 0 (0) | 0.20 |
|  | Day 6 | 1 (1) | 3 (3) | 3 (3) | 0 (0) | 0.27 |
|  | Day 7 | 1 (1) | 3 (3) | 1 (1) | 0 (0) | 0.25 |
| Joint pain | Day of vaccination | 11 (10) | 3 (3) | 10 (9) | 9 (8) | 0.13 |
|  | Day 1 | 24 (23) | 21 (20) | 25 (22) | 20 (18) | 0.80 |
|  | Day 2 | 11 (10) | 4 (4) | 9 (8) | 9 (8) | 0.29 |
|  | Day 3 | 3 (3) | 3 (3) | 3 (3) | 1 (1) | 0.74 |
|  | Day 4 | 2 (2) | 1 (1) | 2 (2) | 1 (1) | 0.86 |
|  | Day 5 | 1 (1) | 0 (0) | 1 (1) | 0 (0) | 0.62 |
|  | Day 6 | 1 (1) | 0 (0) | 1 (1) | 0 (0) | 0.62 |
|  | Day 7 | 1 (1) | 0 (0) | 1 (1) | 0 (0) | 0.62 |
| Redness at vaccination site | Day of vaccination | 4 (4) | 4 (4) | 4 (4) | 3 (3) | 0.97 |
|  | Day 1 | 7 (7) | 6 (6) | 4 (4) | 3 (3) | 0.49 |
|  | Day 2 | 6 (6) | 4 (4) | 3 (3) | 2 (2) | 0.43 |
|  | Day 3 | 4 (4) | 2 (2) | 2 (2) | 2 (2) | 0.78 |
|  | Day 4 | 2 (2) | 2 (2) | 1 (1) | 2 (2) | 0.88 |
|  | Day 5 | 2 (2) | 2 (2) | 1 (1) | 1 (1) | 0.77 |
|  | Day 6 | 1 (1) | 2 (2) | 0 (0) | 1 (1) | 0.47 |
|  | Day 7 | 1 (1) | 2 (2) | 0 (0) | 1 (1) | 0.47 |
| Swelling at vaccination site | Day of vaccination | 5 (5) | 2 (2) | 2 (2) | 2 (2) | 0.52 |
|  | Day 1 | 7 (7) | 5 (5) | 8 (7) | 6 (5) | 0.87 |
|  | Day 2 | 7 (7) | 4 (4) | 6 (5) | 5 (5) | 0.80 |
|  | Day 3 | 4 (4) | 3 (3) | 6 (5) | 2 (2) | 0.51 |
|  | Day 4 | 1 (1) | 3 (3) | 3 (3) | 1 (1) | 0.63 |
|  | Day 5 | 1 (1) | 2 (2) | 3 (3) | 1 (1) | 0.79 |
|  | Day 6 | 1 (1) | 2 (2) | 1 (1) | 1 (1) | 0.88 |
|  | Day 7 | 1 (1) | 1 (1) | 1 (1) | 1 (1) | 1.00 |
| Pain at vaccination site | Day of vaccination | 35 (33) | 46 (43) | 39 (35) | 58 (52) | 0.02 |
|  | Day 1 | 57 (54) | 58 (55) | 65 (58) | 72 (65) | 0.36 |
|  | Day 2 | 35 (33) | 41 (39) | 42 (38) | 50 (45) | 0.36 |
|  | Day 3 | 24 (23) | 20 (19) | 22 (20) | 22 (20) | 0.89 |
|  | Day 4 | 12 (11) | 12 (11) | 8 (7) | 11 (10) | 0.68 |
|  | Day 5 | 6 (6) | 9 (8) | 5 (4) | 5 (5) | 0.58 |
|  | Day 6 | 5 (5) | 7 (7) | 4 (4) | 4 (4) | 0.72 |
|  | Day 7 | 5 (5) | 4 (4) | 3 (3) | 4 (4) | 0.85 |
| Note: Values are number (percentage). P-values are based on Fisher’s exact test. Bold p-values are significant at the 1% alpha level. | | | | | | |

**Supplementary table 3. Timing of side-effects over seven days following boost.**

| In the seven days after your second vaccination did you suffer from one of the following side-effects: | | Ad26.COV2.S/  Ad26.COV2.S | Ad26.COV2.S/  mRNA-1273 | Ad26.COV2.S/  BNT162b2 | P-value |
| --- | --- | --- | --- | --- | --- |
|  |  | N= 106 | N= 112 | N= 111 |  |
| Fatigue | Day of vaccination | 40 (38) | 46 (41) | 29 (26) | 0.05 |
|  | Day 1 | 63 (59) | 90 (80) | 73 (66) | **0.002** |
|  | Day 2 | 34 (32) | 52 (46) | 37 (33) | 0.05 |
|  | Day 3 | 15 (14) | 28 (25) | 19 (17) | 0.11 |
|  | Day 4 | 13 (12) | 13 (12) | 12 (11) | 0.98 |
|  | Day 5 | 10 (9) | 8 (7) | 11 (10) | 0.76 |
|  | Day 6 | 6 (6) | 5 (4) | 8 (7) | 0.67 |
|  | Day 7 | 4 (4) | 2 (2) | 5 (5) | 0.51 |
| Chills | Day of vaccination | 20 (19) | 31 (28) | 9 (8) | **<0.001** |
|  | Day 1 | 18 (17) | 37 (33) | 23 (21) | 0.02 |
|  | Day 2 | 4 (4) | 8 (7) | 6 (5) | 0.62 |
|  | Day 3 | 0 (0) | 1 (1) | 1 (1) | 1.00 |
|  | Day 4 | 0 (0) | 0 (0) | 0 (0) | NA |
|  | Day 5 | 0 (0) | 0 (0) | 2 (2) | 0.21 |
|  | Day 6 | 0 (0) | 1 (1) | 2 (2) | 0.66 |
|  | Day 7 | 0 (0) | 1 (1) | 1 (1) | 1.00 |
| Fever | Day of vaccination | 14 (13) | 20 (18) | 7 (6) | 0.03 |
|  | Day 1 | 17 (16) | 41 (37) | 13 (12) | **<0.001** |
|  | Day 2 | 2 (2) | 10 (9) | 4 (4) | 0.06 |
|  | Day 3 | 1 (1) | 2 (2) | 1 (1) | 1.00 |
|  | Day 4 | 1 (1) | 0 (0) | 1 (1) | 0.55 |
|  | Day 5 | 1 (1) | 0 (0) | 1 (1) | 0.55 |
|  | Day 6 | 0 (0) | 0 (0) | 1 (1) | 0.66 |
|  | Day 7 | 0 (0) | 0 (0) | 1 (1) | 0.66 |
| Nausea | Day of vaccination | 6 (6) | 8 (7) | 4 (4) | 0.49 |
|  | Day 1 | 19 (18) | 21 (19) | 14 (13) | 0.42 |
|  | Day 2 | 6 (6) | 5 (4) | 5 (5) | 0.90 |
|  | Day 3 | 2 (2) | 2 (2) | 1 (1) | 0.87 |
|  | Day 4 | 2 (2) | 0 (0) | 1 (1) | 0.21 |
|  | Day 5 | 0 (0) | 0 (0) | 1 (1) | 0.66 |
|  | Day 6 | 2 (2) | 0 (0) | 0 (0) | 0.10 |
|  | Day 7 | 3 (3) | 1 (1) | 2 (2) | 0.46 |
| Headache | Day of vaccination | 33 (31) | 42 (38) | 20 (18) | **0.004** |
|  | Day 1 | 50 (47) | 72 (64) | 64 (58) | 0.04 |
|  | Day 2 | 22 (21) | 42 (38) | 32 (29) | **0.02** |
|  | Day 3 | 10 (9) | 16 (14) | 19 (17) | 0.24 |
|  | Day 4 | 9 (8) | 6 (5) | 11 (10) | 0.44 |
|  | Day 5 | 5 (5) | 2 (2) | 8 (7) | 0.14 |
|  | Day 6 | 2 (2) | 2 (2) | 7 (6) | 0.12 |
|  | Day 7 | 2 (2) | 0 (0) | 6 (5) | 0.02 |
| Muscle ache | Day of vaccination | 29 (27) | 36 (32) | 26 (23) | 0.35 |
|  | Day 1 | 37 (35) | 67 (60) | 56 (50) | **<0.001** |
|  | Day 2 | 20 (19) | 43 (38) | 31 (28) | **0.006** |
|  | Day 3 | 6 (6) | 12 (11) | 9 (8) | 0.44 |
|  | Day 4 | 4 (4) | 2 (2) | 4 (4) | 0.67 |
|  | Day 5 | 1 (1) | 1 (1) | 1 (1) | 1.00 |
|  | Day 6 | 0 (0) | 0 (0) | 0 (0) | NA |
|  | Day 7 | 0 (0) | 0 (0) | 0 (0) | NA |
| Joint pain | Day of vaccination | 10 (9) | 7 (6) | 7 (6) | 0.60 |
|  | Day 1 | 21 (20) | 30 (27) | 17 (15) | 0.10 |
|  | Day 2 | 14 (13) | 15 (13) | 5 (5) | 0.04 |
|  | Day 3 | 5 (5) | 4 (4) | 0 (0) | 0.06 |
|  | Day 4 | 4 (4) | 1 (1) | 0 (0) | 0.05 |
|  | Day 5 | 1 (1) | 2 (2) | 0 (0) | 0.54 |
|  | Day 6 | 1 (1) | 1 (1) | 1 (1) | 1.00 |
|  | Day 7 | 0 (0) | 1 (1) | 1 (1) | 1.00 |
| Redness at vaccination site | Day of vaccination | 1 (1) | 8 (7) | 1 (1) | **0.01** |
|  | Day 1 | 2 (2) | 13 (12) | 1 (1) | **<0.001** |
|  | Day 2 | 2 (2) | 13 (12) | 0 (0) | **<0.001** |
|  | Day 3 | 2 (2) | 11 (10) | 0 (0) | **<0.001** |
|  | Day 4 | 2 (2) | 7 (6) | 0 (0) | **0.009** |
|  | Day 5 | 1 (1) | 6 (5) | 0 (0) | 0.02 |
|  | Day 6 | 0 (0) | 1 (1) | 0 (0) | 1.00 |
|  | Day 7 | 0 (0) | 1 (1) | 0 (0) | 1.00 |
| Swelling at vaccination site | Day of vaccination | 6 (6) | 15 (13) | 12 (11) | 0.15 |
|  | Day 1 | 6 (6) | 30 (27) | 17 (15) | **<0.001** |
|  | Day 2 | 7 (7) | 25 (22) | 13 (12) | **0.003** |
|  | Day 3 | 1 (1) | 10 (9) | 7 (6) | 0.02 |
|  | Day 4 | 1 (1) | 5 (4) | 3 (3) | 0.31 |
|  | Day 5 | 0 (0) | 3 (3) | 3 (3) | 0.25 |
|  | Day 6 | 0 (0) | 0 (0) | 0 (0) | NA |
|  | Day 7 | 0 (0) | 0 (0) | 0 (0) | NA |
| Pain at vaccination site | Day of vaccination | 56 (53) | 76 (68) | 71 (64) | 0.06 |
|  | Day 1 | 67 (63) | 101 (90) | 95 (86) | **<0.001** |
|  | Day 2 | 49 (46) | 82 (73) | 64 (58) | **<0.001** |
|  | Day 3 | 26 (25) | 44 (39) | 30 (27) | 0.04 |
|  | Day 4 | 14 (13) | 13 (12) | 12 (11) | 0.89 |
|  | Day 5 | 5 (5) | 6 (5) | 3 (3) | 0.62 |
|  | Day 6 | 1 (1) | 2 (2) | 1 (1) | 1.00 |
|  | Day 7 | 1 (1) | 1 (1) | 1 (1) | 1.00 |
| Note: Values are number (percentage). P-values are based on Fisher’s exact test. Bold p-values are significant at the 1% alpha level. | | | | | |

**Supplementary table 4. Data visually presented in Figure 4.**

| In the seven days after your second vaccination did you suffer from one of the following side-effects: | | Ad26.COV2.S/ Ad26.COV2.S | Ad26.COV2.S/ mRNA-1273 | Ad26.COV2.S/ BNT162b2 | p-value |
| --- | --- | --- | --- | --- | --- |
|  |  | N= 106 | N= 111 | N= 111 |  |
| Fatigue | Not at all | 36 (34) | 18 (16) | 30 (27) | 0.07 |
|  | A little, but did not hinder my daily activity | 46 (43) | 54 (48) | 54 (49) |  |
|  | Quite, it hindered my daily activity | 20 (19) | 32 (29) | 23 (21) |  |
|  | Significantly, I could not perform my daily activity | 4 (4) | 8 (7) | 4 (4) |  |
| Chills | Not at all | 78 (74) | 56 (50) | 81 (73) | **<0.001** |
|  | A little, but did not hinder my daily activity | 22 (21) | 34 (30) | 24 (22) |  |
|  | Quite, it hindered my daily activity | 6 (6) | 15 (13) | 6 (5) |  |
|  | Significantly, I could not perform my daily activity | 0 (0) | 7 (6) | 0 (0) |  |
| Fever | Not at all | 84 (79) | 69 (62) | 93 (84) | **0.003** |
|  | 38.0-38.4 | 17 (16) | 28 (25) | 15 (14) |  |
|  | 38.5-38.9 | 4 (4) | 12 (11) | 3 (3) |  |
|  | >39.0 | 1 (1) | 3 (3) | 0 (0) |  |
| Nausea | Not at all | 83 (78) | 89 (79) | 91 (82) | 0.60 |
|  | A little, but did not hinder my daily activity | 18 (17) | 19 (17) | 19 (17) |  |
|  | Quite, it hindered my daily activity | 5 (5) | 3 (3) | 1 (1) |  |
|  | Significantly, I could not perform my daily activity | 0 (0) | 1 (1) | 0 (0) |  |
| Headache | Not at all | 44 (42) | 28 (25) | 41 (37) | 0.17 |
|  | A little, but did not hinder my daily activity | 44 (42) | 56 (50) | 45 (41) |  |
|  | Quite, it hindered my daily activity | 17 (16) | 25 (22) | 21 (19) |  |
|  | Significantly, I could not perform my daily activity | 1 (1) | 3 (3) | 4 (4) |  |
| Muscle ache | Not at all | 59 (56) | 42 (38) | 52 (47) | **0.003** |
|  | A little, but did not hinder my daily activity | 42 (40) | 47 (42) | 51 (46) |  |
|  | Quite, it hindered my daily activity | 5 (5) | 21 (19) | 8 (7) |  |
|  | Significantly, I could not perform my daily activity | 0 (0) | 2 (2) | 0 (00 |  |
| Joint pain | Not at all | 80 (75) | 80 (71) | 90 (81) | 0.57 |
|  | A little, but did not hinder my daily activity | 20 (19) | 26 (23) | 17 (15) |  |
|  | Quite, it hindered my daily activity | 6 (6) | 5 (4) | 4 (4) |  |
|  | Significantly, I could not perform my daily activity | 0 (0) | 1 (1) | 0 (0) |  |
| Red spot at vaccination site | Not at all | 103 (97) | 94 (84) | 109 (98) | **<0.001** |
|  | 2.5-5.0 cm | 3 (3) | 13 (12) | 2 (2) |  |
|  | 5.1-10.0 cm | 0 (0) | 4 (4) | 0 (0) |  |
|  | >10.0 cm | 0 (0) | 1 (1) | 0 (0) |  |
| Swelling at vaccination site | Not at all | 96 (91) | 79 (71) | 91 (82) | **0.003** |
|  | 2.5-5.0 cm | 10 (9) | 28 (25) | 17 (15) |  |
|  | 5.1-10.0 cm | 0 (0) | 4 (4) | 3 (3) |  |
|  | >10.0 cm | 0 (0) | 1 (1) | 0 (0) |  |
| Pain at vaccination site | Not at all | 33 (31) | 9 (8) | 14 (13) | **<0.001** |
|  | A little, but did not hinder my daily activity | 71 (67) | 74 (66) | 83 (75) |  |
|  | Quite, it hindered my daily activity and needed pain killers (e.g., paracetamol) | 2 (2) | 28 (25) | 14 (13) |  |
|  | Significantly, I could not perform my daily activity and needed pain killers (e.g., paracetamol) | 0 (0) | 1 (1) | 0 (0) |  |
| Note: Values are number (percentage). P-values are based on Fisher’s exact test. Bold p-values are significant at the 1% alpha level. | | | | | |

**References**

1.	Bryan A, Pepper G, Wener MH, et al. Performance Characteristics of the Abbott Architect SARS-CoV-2 IgG Assay and Seroprevalence in Boise, Idaho. J Clin Microbiol 2020;58.

2.	Okba NMA, Muller MA, Li W, et al. Severe Acute Respiratory Syndrome Coronavirus 2-Specific Antibody Responses in Coronavirus Disease Patients. Emerg Infect Dis 2020;26:1478-88.

3.	Geers D, Shamier MC, Bogers S, et al. SARS-CoV-2 variants of concern partially escape humoral but not T-cell responses in COVID-19 convalescent donors and vaccinees. Science Immunology 2021.

4.	Sanders J-S, Bemelman FJ, Messchendorp AL, et al. The RECOVAC Immune-Response Study The immunogenicity, tolerability and safety of COVID-19 vaccination in patients with chronic kidney disease, on dialysis, or living with a kidney transplant. Transplantation 2021;forthcoming.

5.	Liu X, Shaw RH, Stuart ASV, et al. Safety and immunogenicity of heterologous versus homologous prime-boost schedules with an adenoviral vectored and mRNA COVID-19 vaccine (Com-COV): a single-blind, randomised, non-inferiority trial. The Lancet 2021;398:856-69.

6.	Borobia AM, Carcas AJ, Pérez-Olmeda M, et al. Immunogenicity and reactogenicity of BNT162b2 booster in ChAdOx1-S-primed participants (CombiVacS): a multicentre, open-label, randomised, controlled, phase 2 trial. The Lancet 2021;398:121-30.

7.	Sablerolles RSG, Goorhuis A, GeurtsvanKessel C, et al. Heterologous Ad26.COV2.S Prime and mRNA-Based Boost COVID-19 Vaccination Regimens: The SWITCH Trial Protocol. Front Immunol 2021.
